# Polygenic risk scores based on European GWAS correlate to disease prevalence differences around the world

**DOI:** 10.1101/2021.11.13.21265898

**Authors:** Pritesh R Jain, Myson Burch, Melanie Martinez, Pablo Mir, Jakub Fichna, Cezary Zekanowski, Renata Rizzo, Zeynep Tümer, Csaba Barta, Evangelia Yannaki, John Stamatoyannopoulos, Petros Drineas, Peristera Paschou

## Abstract

**Background:** Complex disorders are caused by a combination of genetic, environmental and lifestyle factors, and their prevalence can vary greatly across different populations. Genome wide association studies (GWAS) can help identify common variants that underlie disease risk. However, despite their increasing number, the vast majority of studies focuses on European populations, leading to questions regarding the transferability of findings to non-Europeans. Here, we investigated whether polygenic risk scores (PRS) based on European GWAS correlate to disease prevalence within Europe and around the world.

**Results:** GWAS summary statistics of 20 different disorders were used to estimate PRS in nine European and 24 worldwide reference populations. We estimated the correlation between average genetic risk for each of the 20 disorders and their prevalence in Europe and around the world. A clear variation in genetic risk was observed based on ancestry and we identified populations that have a higher genetic liability for developing certain disorders both within European and global regions. We also found significant correlations between worldwide disease prevalence and PRS for 13 of the studied disorders with obesity genetic risk having the highest correlation to disease prevalence. For these 13 disorders we also found that the loci used in PRS are significantly more conserved across the different populations compared to randomly selected SNPs as revealed by Fst and linkage disequilibrium structure.

**Conclusion:** Our results show that PRS of world populations calculated based on European GWAS data can significantly capture differences in disease risk and identify populations with the highest genetic liability to develop various conditions. Our findings point to the potential transferability of European-based GWAS results to non-European populations and provide further support for the validity of GWAS.

## Background

Complex disorders are caused by the interaction of genetic, environmental and lifestyle factors. Most disorders that are frequent in the human populations fall under this category (1) and their prevalence varies greatly around the world (2). Understanding the basis of this prevalence difference can help disentangle the interaction among different factors causing complex disorders and identify groups of people who may be at a greater risk of developing certain disorders. This could become the basis of the implementation of early intervention strategies for populations at higher risk with significant benefits for public health.

The genetic component underlying complex disorders is not easy to quantify. It is highly polygenic in nature, possibly involving hundreds of genetic variants each with a very small effect on disease liability and occurrence (3). To measure the genetic risk of developing a specific disorder, it is possible to combine the effects of genomewide individual variants deriving a polygenic risk score (PRS) to quantify the genetic liability of a disorder and compare the risk of developing complex disorders across various populations (4). PRS of an individual for a specific disorder is estimated by the sum of the number of risk alleles weighted by the effect size of a specific allele (5) which is obtained from genomewide association studies (GWAS). With the availability of large-scale datasets, thousands of GWAS have been performed for various traits and conditions thus providing a large database of effect sizes that can be used to estimate PRS for different complex disorders (6).

PRS has become an increasingly powerful tool to identify individuals at higher risk of developing complex disorders and could help explain the proportion of genetic variance that seems to be missing when focusing only on genome-wide significant hits (7,8). However, to date, PRS-based research has been hampered by the lack of GWAS summary statistics data from diverse populations. It was recently highlighted that about 70% of GWAS studies since 2008 have used samples solely from European populations (9). Previous studies have shown that the predictive power of PRS based on European GWAS is comparatively poorer in non-Europeans and this decline increases with divergence from European genetic structure (10). The loss in prediction accuracy could be due to linkage disequilibrium (LD) structure and allele frequency differences between populations which in turn could lead to differences in the effect size estimates from the GWAS based on one population compared to another (10–12). Overall, it is recommended to avoid cross-population PRS calculation and to use GWAS data from populations with similar population structure for the estimation of genetic risk scores,

At the same time, systematic studies attempting to evaluate the degree to which PRS can predict disease prevalence in different populations have not been performed to date in Europeans or non-Europeans. If such correlation of PRS to epidemiology exists, it would significantly boost confidence in the validity of GWAS results and the potential for their use as a tool in the design of public health studies. In the case of non-Europeans, based on the above-mentioned observations and known differences in LD structure around the world, one would expect that such correlations would be very poor. Here, we embark on a systematic exploration of the genetic architecture of 20 different complex disorders, using European GWAS to estimate average genetic risk within Europe but also around the world. Intriguingly, we find that PRS correlates to disease prevalence difference around the world for multiple disorders and show that this correlation might be explained by conservation of genetic regions that have been implicated in disease susceptibility via GWAS. Our study highlights the value of GWAS results and the potential of the use of PRS to identify high-risk populations around the world.

## Results

### PRS of complex disorders in European Populations

We began by exploring PRS across a dataset of nine different European populations (2,109 individuals) (supplementary table 1) for 20 different complex disorders for which recent large GWAS were available (see methods and supplementary table 2) (13–31)). Studied disorders can be grouped under five general categories (cardiovascular, neurological, autoimmune, metabolic, and psychiatric). As expected, principal component analysis (PCA) showed that the analyzed samples clustered based on their geography (Figure 1a). PRS was calculated using PRSice-2 and a simple clumping and p-value thresholding-based method to correct for LD and select SNPs used to estimate PRS (32). LD clumping was performed separately for each ancestral group with a threshold of r^2^=0.1 within a 250kb distance and six p-value thresholds (5×10^−8^, 5×10^−5^, 0.001, 0.01, 0.05 and 1) were used for this analysis. The final number of SNPs used at each threshold are shown in additional file 1.

**Figure 1:**
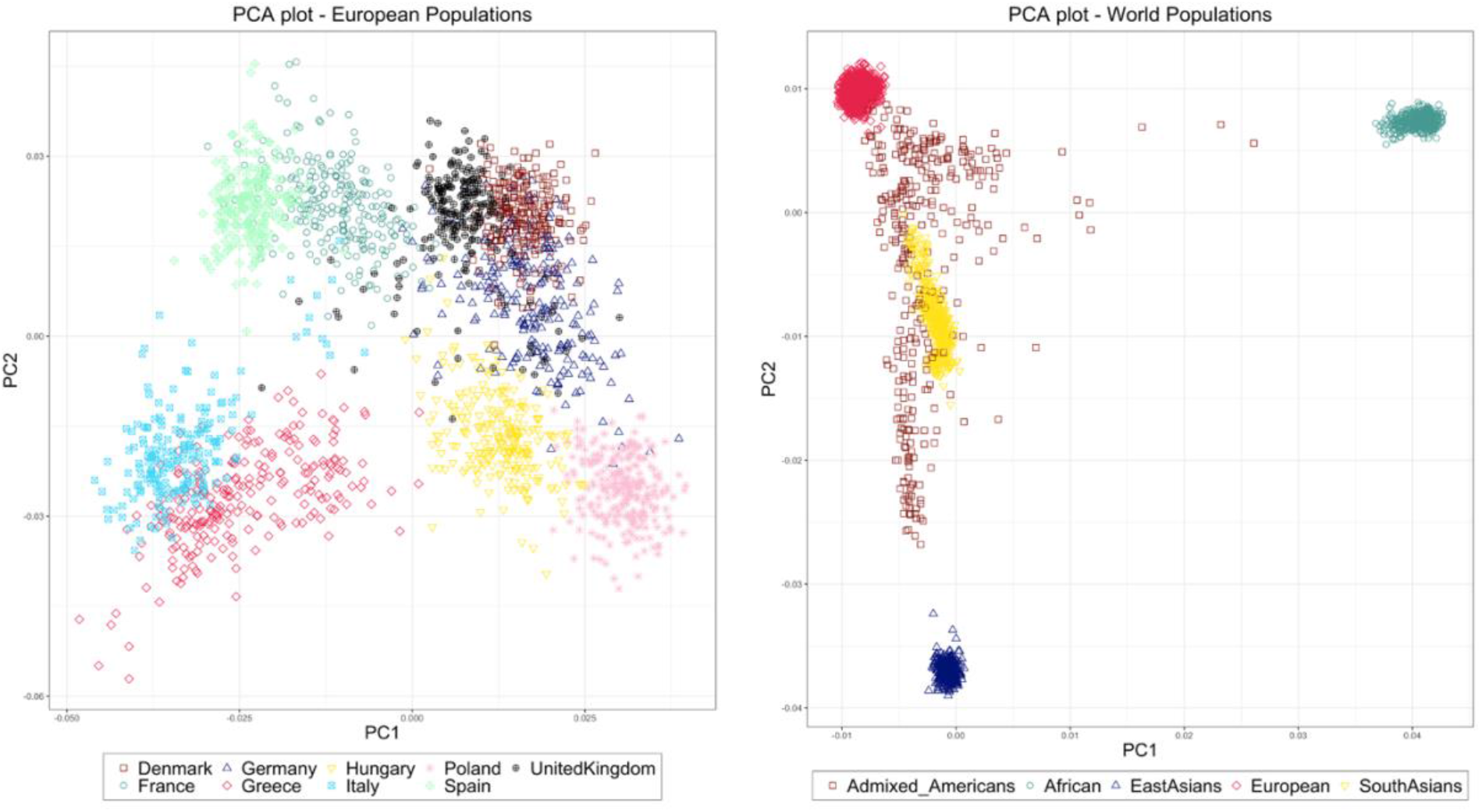
PCA plot of European and World Populations. The left panel shows distribution of 2,109 European samples based on top two PCs colored and shaped based on their country of origin. The right panel shows the distribution of 3,953 global samples based on top 2 PCs colored and shaped based on their region/ethnicity.

Figure 2 shows the overall results at threshold p-value<5×10^−5^. Individuals from southern European countries (Greece and Italy) were found to carry 1.5 – 2.7 times lower risk of developing most autoimmune disorders and 1.7 – 2.2 times higher risk of developing metabolic disorders compared to central and northern European populations. The highest risk for coronary artery disease (CAD) was observed in populations from central European countries like Hungary and Poland. These populations also showed two times higher genetic risk for Parkinson disease (PD) compared to other Europeans in this analysis. By contrast, we found that individuals from northern European countries like Denmark and the United Kingdom (UK) have lower risk for neurological disorders and higher risk for autoimmune diseases. Among the different psychiatric disorders that we analyzed, the highest PRS for autism spectrum disorders (ASD) and major depressive disorder (MDD) was observed in the Greek and Italian populations and the lowest PRS was seen in the Polish population, which inversely has the highest genetic risk for anxiety disorders (ANX), schizophrenia (SCZ) and bipolar disorder (BPD). The overall genetic risk of psychiatric disorders is lower in Northern European populations, except for ADHD for which Denmark has the highest risk score estimate among all the analyzed groups.

**Figure 2:**
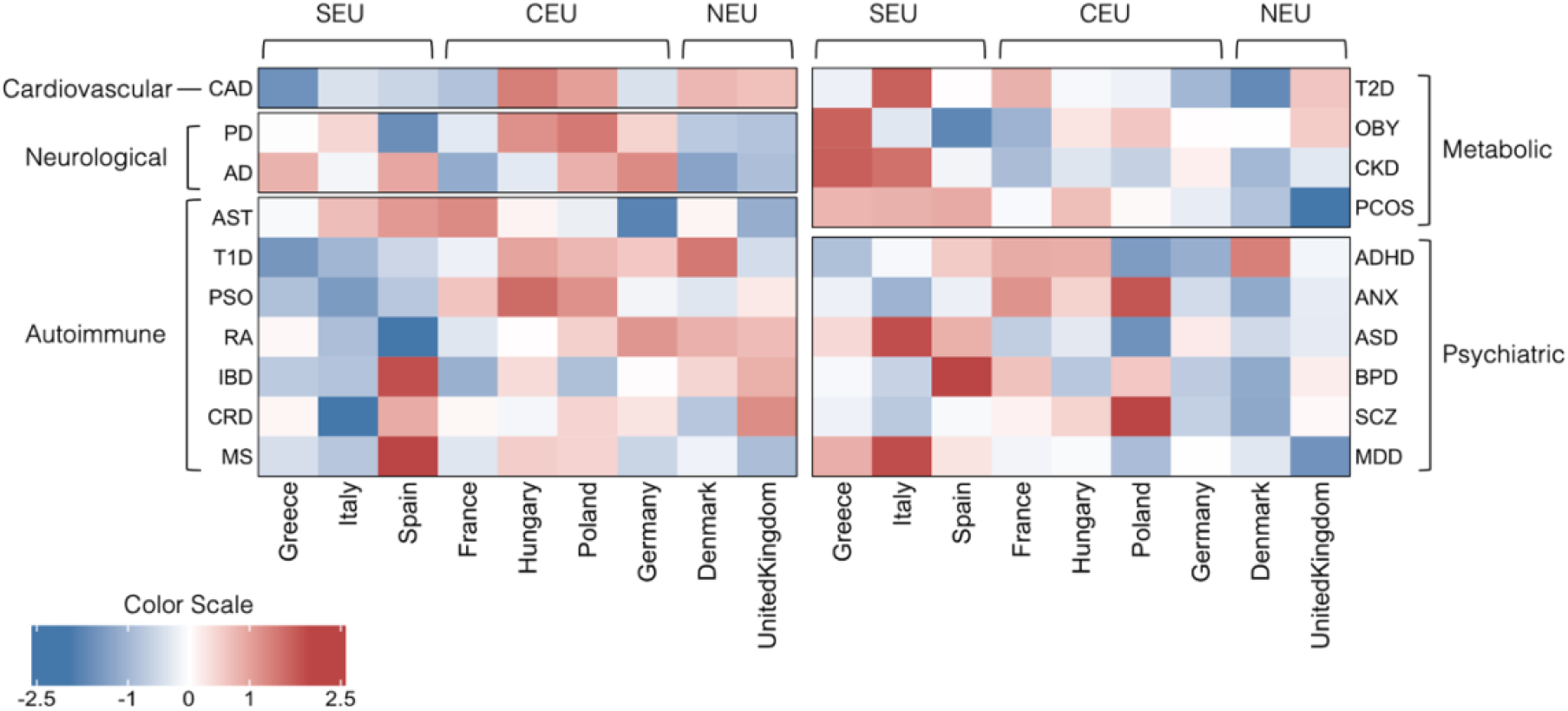
Heatmap of Average PRS Scores (p-value<5×10^−5^) of 20 Disorders across European Populations. The disorders are grouped based on the disease domain and the Populations are arranged based on their geographical location going from southern to northern countries. Shades of cells indicate the standardized avg. genetic risk of each disorder for each population. A higher risk is shown by red and lower risk is indicated by blue [SEU – South Europeans, CEU – Central Europeans, NEU – North Europeans.]

Results for PRS calculated at other p-value thresholds revealed an overall similar distribution of disease risk in different populations. At higher p-value thresholds, the differences between populations became more distinct and stronger clustering was observed between countries in the same region (supplementary figure 1 and additional file 2)

### Genetic Risk in Global Populations

We continued to explore the use of European-GWAS PRS, expanding our analysis to global populations (1000 genomes phase 3 data (33)). The data set is made up of 3,953 individuals from 24 different populations in five regions of the world – Africans (AFR), South Asians (SAS), East Asians (EAS) and Admixed Americans (AMR) (supplementary table 1). The PCA plot of the global data again showed that the populations are very tightly clustered based on their regions of origin except for the AMR samples which are distributed along a cline (fig 1b).

Again, PRS scores were calculated for the global populations at six thresholds (additional file 3). In Figure 3, we compared the average PRS calculated at a threshold of p-value<5×10^−5^ for 24 populations from five regions of the world. Genetic risk for the different studied disorders was observed to follow a pattern of distribution reminiscent of geography. Populations originating from the same region mostly tend to have a uniform genetic risk score as compared to risk between populations from different regions.

**Figure 3:**
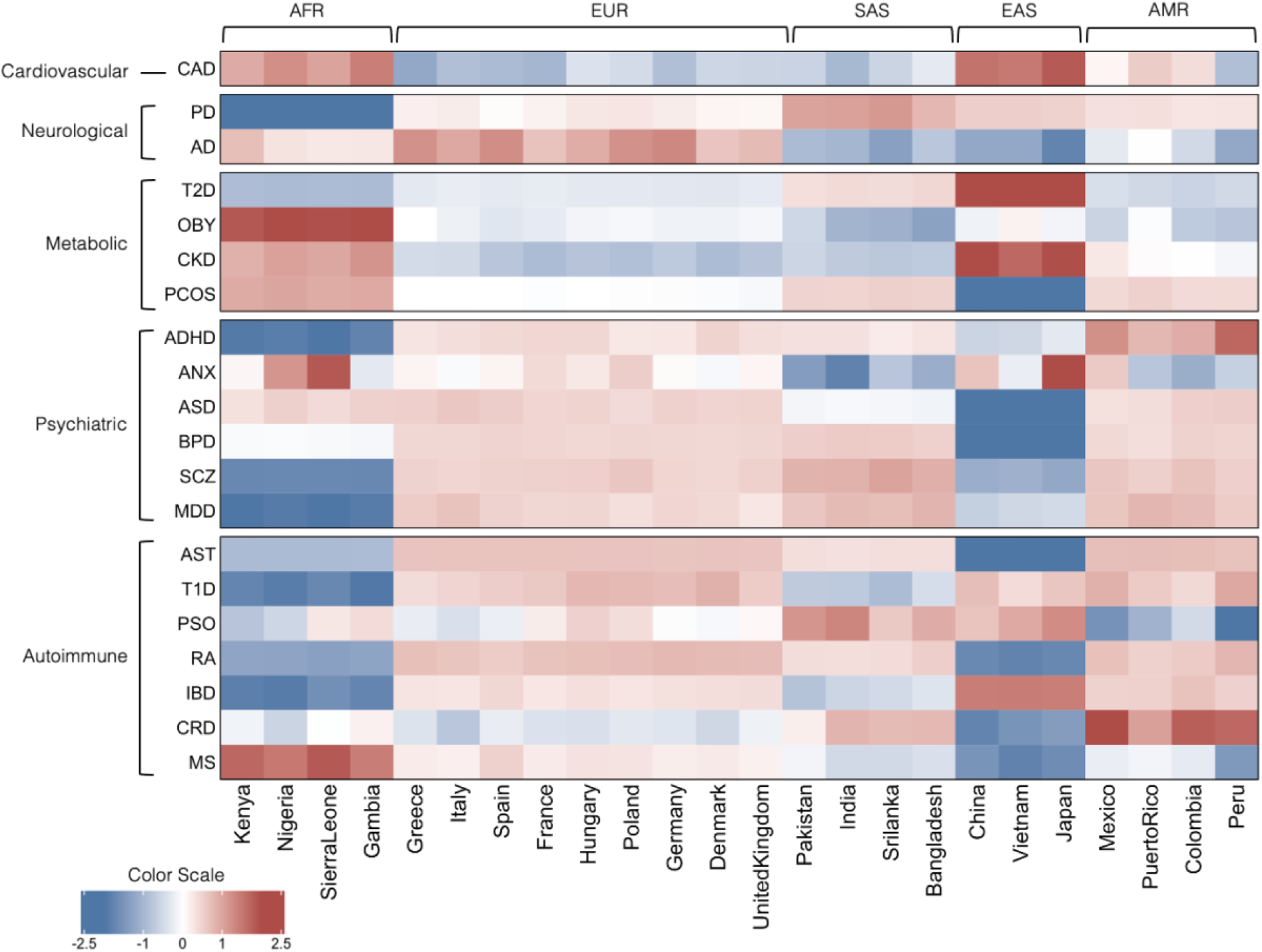
Heatmap of Average PRS Scores (p-value<5×10^−5^) of 20 Disorders across World Populations. The disorders are grouped based on the disease domain and the populations are arranged based on their geographical location and ancestry starting with Africans (AFR) and followed by Europeans (EUR), South Asians (SAS), East Asians (EAS) and Admixed Americans (AMR) Shades of cells indicate the standardized avg. genetic risk of each disorder for each population. A higher risk is shown by red and lower risk is indicated by blue.

African populations were found to have a 1.3 – 2.5 times higher PRS for obesity (OBY) and polycystic ovary syndrome (PCOS) compared to non-African populations. These populations also had 2.2 times lower risk of type 1 diabetes (T1D) and inflammatory bowel disease (IBD); and around 1.9 – 2.4 times lower risk scores for psychiatric disorders like ADHD, SCZ and MDD compared to the rest of the populations. East Asian populations were found to carry 1.8 – 2.3 times lower risk scores for autoimmune conditions like asthma (AST), multiple sclerosis (MS), rheumatoid arthritis (RA) and IBD; and 2.8 times lower PRS for psychiatric disorders like ASD and BPD compared to the other regions. They also had the highest risk for CAD, type 2 diabetes (T2D) and chronic kidney disease (CKD) which was 1.7 – 2.6 times greater than the other populations. Europeans were found to carry a 1.2 – 1.6 times higher risk of Alzheimer disease (AD) and moderately high risk for most autoimmune and psychiatric diseases compared to non-European populations. The lowest genetic risk for European individuals was observed for metabolic and cardiovascular disorders. South Asians followed a distribution pattern intermediate to that observed for East Asians and Europeans. Along with their East Asians neighbors, populations from countries like India, Pakistan, Bangladesh and Srilanka were found to have a higher risk of T2D, and the lowest risk of obesity. They also had the highest PRS for PD and psoriasis (PSO) which was 1.3 times higher than the other populations. The AMR populations were found to carry the highest risk for ADHD which was 1.6 times higher than the rest of the populations and had risk estimates similar to Europeans for other psychiatric and metabolic disorders.

We examined the distribution of PRS calculated at other p-value thresholds and observed that the genetic risk distribution for certain disorders changes at different thresholds. The most significant changes are observed for T2D, OBY, ANX and MS at threshold p-value<1 (supplementary figure 2). The std. average scores at all thresholds are listed in additional file 3.

### Correlation between PRS and prevalence of complex disorders around the world

To determine whether the average PRS of the studied complex disorders could partly explain disease prevalence within Europe, we calculated the correlation between these measures. To test for statistical significance, we calculated an empirical p-value for significant associations based on random SNP sets as explained in the methods. Results are shown in table 1 and supplementary table 3. We observed significant correlation between prevalence and PRS at different thresholds for seven disorders. We proceeded to explore the potential correlation of PRS based on European GWAS to disease prevalence in non-European populations. The mean prevalence of the disorders across the five ancestral populations that we studied is shown in figure 4, the prevalence of each country is shown in additional file 4, and the results of the correlation analysis are shown in table 2 and supplementary table 4. Of the 20 analyzed traits, we found significant correlation between the prevalence of 13 disorders with PRS calculated for at least one p-value threshold. We observed a significant correlation of PRS to disease prevalence for all studied autoimmune disorders except psoriasis with the strongest correlation seen for CRD (R^2^ = 0.6, p=0.001) and PRS calculated at a threshold of p-value<0.01. Among metabolic conditions, significant correlations between PRS and disease prevalence were observed for obesity (R^2^ = 0.73, p=0.001) and T2D (R^2^ = 0.55, p=0.003) PRS at the threshold of p-value< 0.001. The average PRS of PD at p-value<5×10^−5^ also had a significant genetic correlation with the prevalence of PD in different populations (R^2^ = 0.39, p=0.026). Among psychiatric disorders, the average PRS at p<5×10^−5^ had a significant correlation to worldwide prevalence of ADHD (R^2^=0.46, p=0.008), BPD (R^2^ = 0.42, p=0.013) and SCZ (R^2^ = 0.45, p=0.011).

**Table 1:**
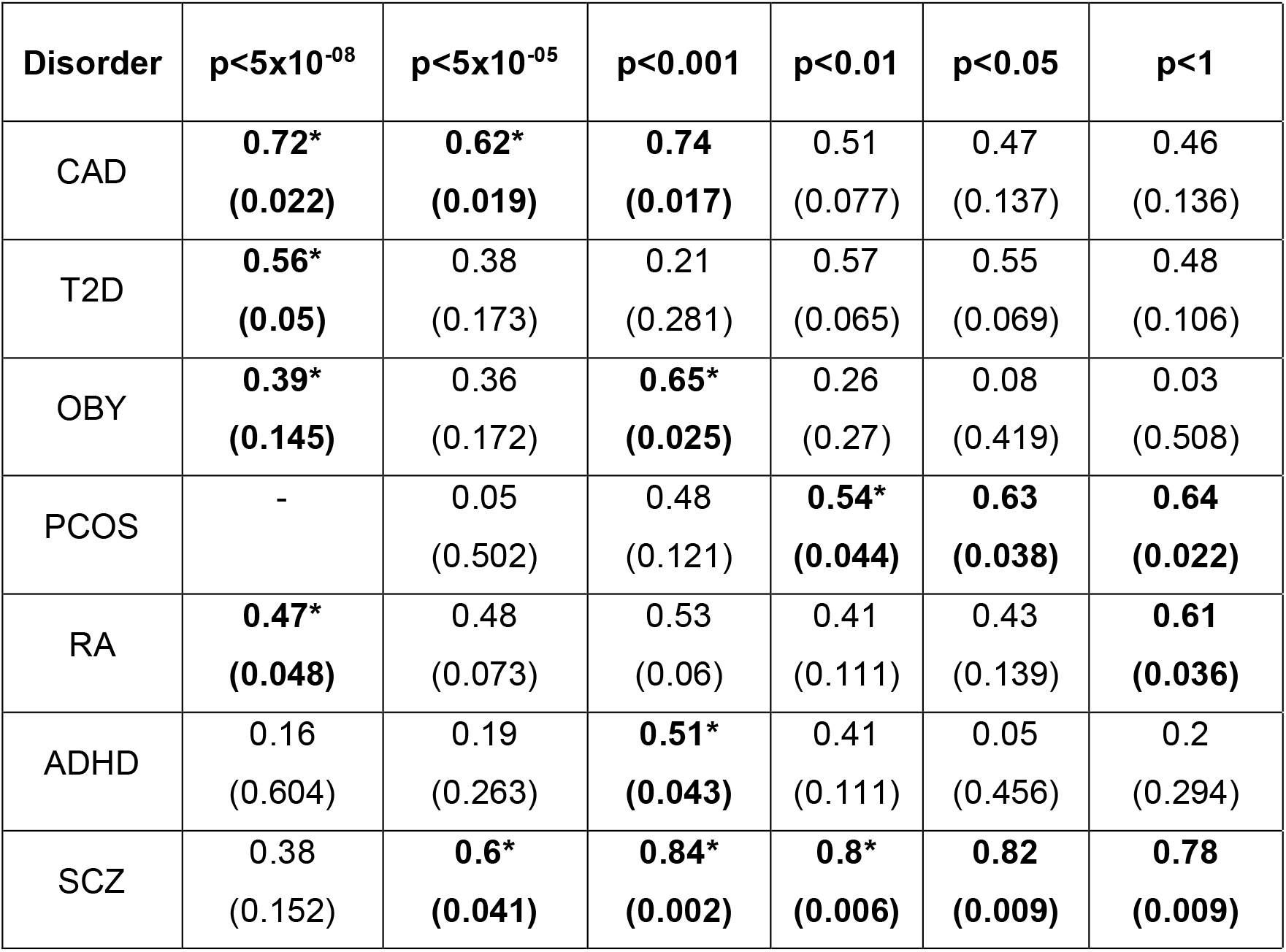
Significant Pearson’s correlations estimate for Average genetic risk of complex disorders and their prevalence in 24 countries. The column headers indicate the p-value threshold for PRS calculation. The value in each cell represents correlation coefficient & p-value based on 1000 permutations (shown in parentheses). The **(*)** indicates empirical p-value<0.05 (based on statistical test)

| Disorder | p<5x10<sup>-08</sup> | p<5x10<sup>-05</sup> | p<0.001 | p<0.01 | p<0.05 | p<1 |
| --- | --- | --- | --- | --- | --- | --- |
| CAD | **0.72\***<br>**(0.022)** | **0.62\***<br>**(0.019)** | **0.74**<br>**(0.017)** | 0.51<br>(0.077) | 0.47<br>(0.137) | 0.46<br>(0.136) |
| T2D | **0.56\***<br>**(0.05)** | 0.38<br>(0.173) | 0.21<br>(0.281) | 0.57<br>(0.065) | 0.55<br>(0.069) | 0.48<br>(0.106) |
| OBY | **0.39\***<br>**(0.145)** | 0.36<br>(0.172) | **0.65\***<br>**(0.025)** | 0.26<br>(0.27) | 0.08<br>(0.419) | 0.03<br>(0.508) |
| PCOS | - | 0.05<br>(0.502) | 0.48<br>(0.121) | **0.54\***<br>**(0.044)** | **0.63**<br>**(0.038)** | **0.64**<br>**(0.022)** |
| RA | **0.47\***<br>**(0.048)** | 0.48<br>(0.073) | 0.53<br>(0.06) | 0.41<br>(0.111) | 0.43<br>(0.139) | **0.61**<br>**(0.036)** |
| ADHD | 0.16<br>(0.604) | 0.19<br>(0.263) | **0.51\***<br>**(0.043)** | 0.41<br>(0.111) | 0.05<br>(0.456) | 0.2<br>(0.294) |
| SCZ | 0.38<br>(0.152) | **0.6\***<br>**(0.041)** | **0.84\***<br>**(0.002)** | **0.8\***<br>**(0.006)** | **0.82**<br>**(0.009)** | **0.78**<br>**(0.009)** |

**Table 2:**
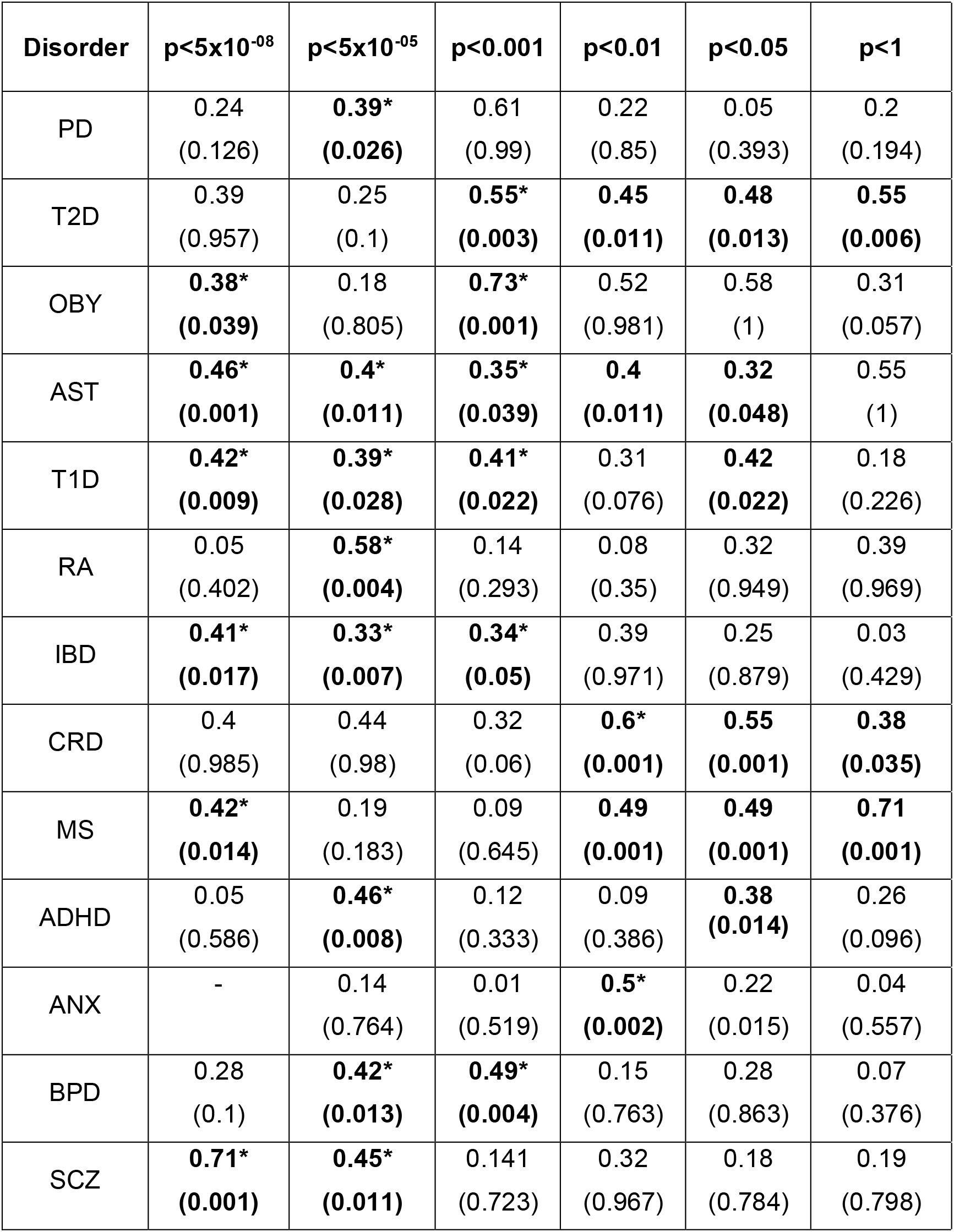
Significant Pearson’s correlations estimate for average genetic risk of complex disorders and their prevalence in 24 countries. The column headers indicate the p-value threshold for PRS calculation. The value in each cell represents correlation coefficient & p-value based on 1000 permutations (shown in parentheses). The **(*)** indicates empirical p-value<0.05 (based on statistical test)

**Figure 4:**
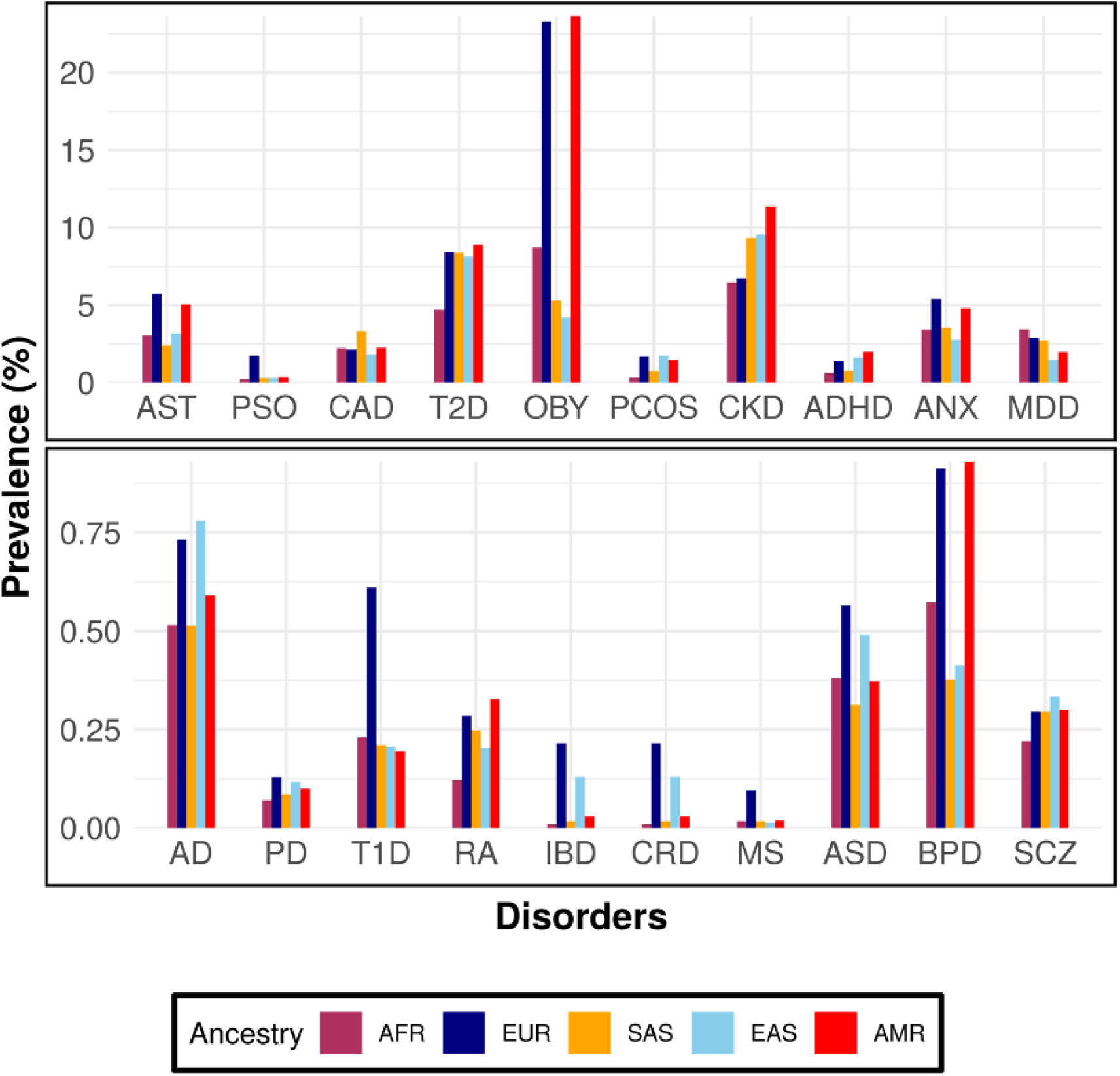
Bar plot showing the mean prevalence of 20 disorders across 5 ancestral groups. The x-axis indicates the ancestral group starting with Africans (AFR) and followed by Europeans (EUR), South Asians (SAS), East Asians (EAS) and Admixed Americans (AMR). The y-axis is the mean prevalence (%) of each group calculated based on the different nationalities in each group.

Next, we investigated whether disorders for which we found significant correlation between prevalence and PRS also had high SNP heritability using LD score regression (LDSC) (34). Results showed that autoimmune disorders except MS had a high SNP heritability ranging from 0.14 – 0.86. Psychiatric disorders like ADHD, BPD and SCZ which have significant correlations with prevalence had heritability estimates 0.23 and 0.3 and 0.157 respectively. PD, AD, and most metabolic disorders showed lower estimates of SNP heritability with less than 5% of variance explained (supplementary table 5). Although there was a positive relationship between the heritability estimates from LDSC and the correlations between PRS and prevalence, no significant associations were observed (supplementary table 6).

### Genetic architecture of disease associated regions used for PRS analysis

The fact that we observed a significant association between worldwide disease prevalence and PRS calculated using European GWAS was quite unexpected. We hypothesized that this could be at least in part due to similarities in the genetic architecture across studied genomic regions around the world. To test this hypothesis, we explored the worldwide structure and allele frequency differences of genomic regions used in our PRS analysis: First, we calculated r^2^(35) for all pairs of variants within 100 kb of the PRS SNPs and performed pairwise comparisons between Europeans and individuals from other geographic regions. Second, we calculated the mean F_ST_ of the PRS SNPs, again performing pairwise comparisons between Europeans and other populations (36). The empirical p-value was calculated using a statistical test based on random SNP sets as explained in the methods section.

Results of our r^2^ analysis showed that for multiple studied disorders, the genetic regions used in PRS calculation show similar LD structure around the world compared to randomly selected regions (empirical p-value <0.05) (figure 5, supplementary table 7). For instance, the regions around the genome-wide significant SNPs used for OBY- and MS-PRS revealed similar LD patterns across all populations indicating that the associated loci have similar genetic structure across all populations. We saw similar LD structure between AFR and European individuals for regions used in PRS estimations for all autoimmune disorders except RA. The LD structure for regions used for PRS in SAS was significantly correlated to European structure for 6 disorders that included OBY, T1D, IBD, MS, ADHD and SCZ. East Asians and Europeans differed mostly with only 3 disorders having significant correlations. Finally, the comparison of LD structure between AMR and Europeans for the studied genetic regions showed significant correlation for almost all disorders.

**Figure 5:**
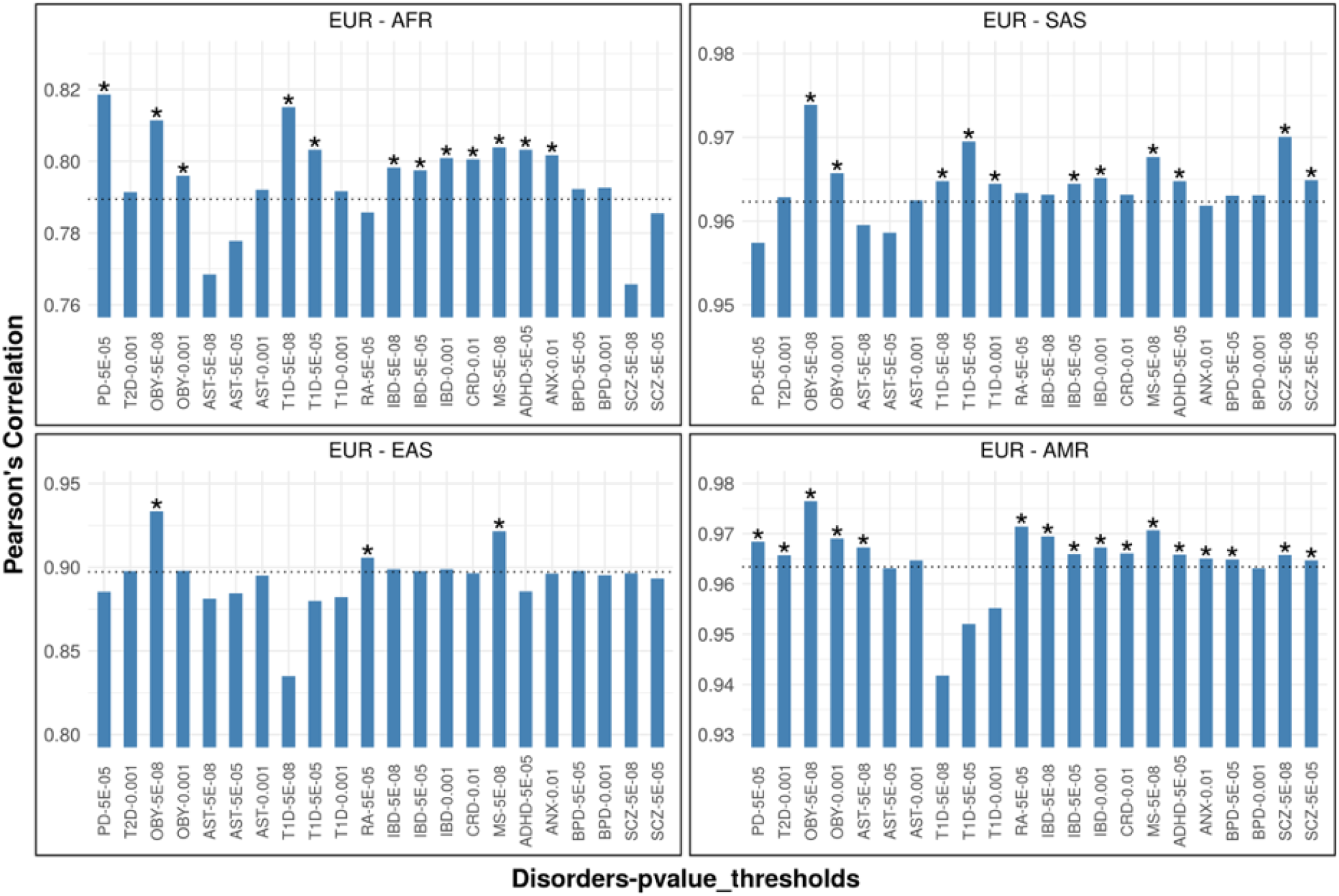
Bar plot showing Pearson’s correlation estimates of r^2^ between four pairs of populations. The x-axis indicates the disorder and the p-value threshold from which the SNPs were selected, and y axis is the correlation coefficient between each pair of populations. The dotted line shows the mean correlation value of the distribution based on 100 Random SNP sets. The (*) indicates empirical p-value < 0.05.

F_ST_ analysis also revealed low genetic differentiation around the world for multiple of the genetic regions used for PRS estimations (figure 6, supplementary table 8). For instance, the SNPs used for PRS calculations of OBY, AST, T1D, IBD, CRD, MS, and ANX had a significantly lower F_ST_ between Africans and Europeans (empirical p-value <0.05). Same as what we observed in the LD analysis, East Asians were often differentiated to Europeans. The results of F_ST_ comparisons of SAS and AMR populations with Europeans were concordant with the results of the LD analysis although fewer significant outcomes were observed for both the populations.

**Figure 6:**
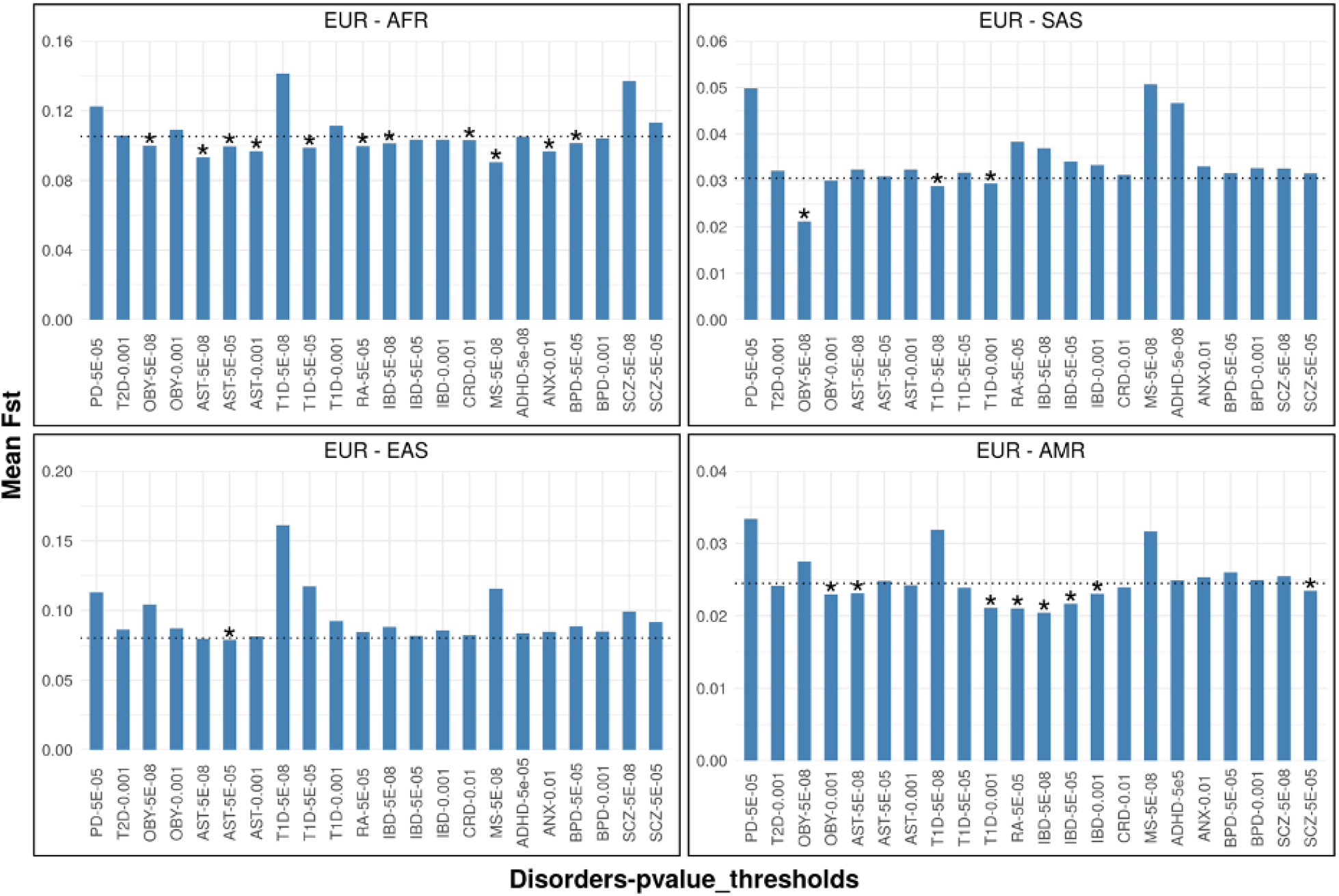
Bar plot showing the mean F_ST_ between four pairs of populations. The x-axis indicates the disorder and the p-value threshold from which the SNPs were selected, and y axis is the mean F_ST_ of each pair of populations. The dotted line shows the mean F_ST_ value of the distribution based on 100 Random SNP sets. The (*) indicates empirical p-value < 0.05.

## Discussion

The prevalence of complex disorders across different populations is quite varied. This may be attributed to a combination of differences in genetic factors, lifestyle, and environment. Here, we explored the genetic component of this variation using PRS to determine and compare the average genetic risk of 20 disorders in individuals belonging to different populations from around the world. Analysis was focused on SNPs that were associated with disease based on European GWAS. Literature describing non-European GWAS is still scarce, thus there is a need to continue to evaluate the transferability of European-based findings to non-European populations as the genetics community works to address this gap through additional studies. As expected, we observed clear differences in the distribution of the average PRS estimates based on ancestry. Most unexpectedly however, for 13 out of the 20 studied disorders, we also showed significant correlation between the average PRS and disease prevalence, a result that indicates that European-based GWAS findings may in fact carry significant value for interpretation of genetic risk in non-European populations.

The differences that we observed in the genetic risk of various disorders could explain greater frequency of specific disorders in different populations. For instance, the highest number of individuals with T2D in the world is reported to be in Asia. Here, we showed that the increased prevalence of this disorder is also accompanied by increased genetic risk for T2D (37). It is also interesting that in Asian populations we found the genetic risk for obesity to be quite low which could explain the unique clinical presentation of diabetic phenotype in Asian populations with lower rates of obesity (38). In a similar fashion, individuals of African ancestry were observed to have a greater genetic risk for developing multiple metabolic disorders, Alzheimer disease, and CAD. Indeed, the prevalence of CAD has been reported to be higher in African Americans than Europeans, and studies have shown that African Americans have the highest risk of being diagnosed with AD and CAD (39–42). Epidemiological studies have also shown that Europeans have the highest lifetime prevalence of mental health conditions. In concordance with this observation, we found higher genetic risk of developing psychiatric conditions in European populations compared to people of other ancestry (43).

The lack of non-European GWAS for the studied disorders is a limitation of this analysis, since there could be a decrease in prediction accuracy of PRS as previously described (9,11,44). However, despite this limitation, we found that average PRS of various non-European populations calculated using GWAS based on Europeans can actually capture differences in disease prevalence across these populations. Intriguingly, we also showed low differentiation of LD structure and allele frequency for SNPs used in PRS calculations suggesting that GWAS may be identifying disease-causing loci that may be conserved across various populations.

Using European GWAS as the basis of this analysis we were able to capture genetic risk differences and correlations to disease prevalence around the world. However, we may have also ignored variants that might be significantly associated with disease in non-European populations (10,45). Another limitation of our analysis is that the awareness regarding various conditions, especially psychiatric disorders, may be low in developing countries and hence the prevalence data might be biased (46,47). This could also explain why not many significant associations were observed for psychiatric disorders in non-European populations.

Identification of populations that carry increased genetic susceptibility to disease could help inform clinical practice and public health strategies. Currently, non-European GWAS are scarce and our knowledge on the genetic architecture of complex disease relies almost solely on the analysis of European genomes. Here, we showed that disease loci identified via European GWAS may have a conserved structure around the world which could explain the correlation of PRS to worldwide disease prevalence that we also observed. However, future studies that include GWAS data based on trans-ancestral populations and use methods that can better adjust for differences in ancestry in base and target datasets by either modelling the LD structure or including annotation and fine-mapping data would improve the prediction accuracy of the risk estimate even further (48–50). Ultimately, combining genetic risk along with information on lifestyle and environmental factors will help fully explain differences in disease prevalence around the world and inform the design of future public health strategies.

## Conclusion

We estimated the genetic risk of 20 complex disorders across five different continental regions to explore whether genetics might help explain disease prevalence distribution around the world. We found that PRS of world populations calculated based on European GWAS data can capture differences in disease risk and could thus be used to identify populations with the highest genetic liability to develop various disorders. Significant correlations were observed between genetic risk and disease prevalence for 13 disorders in different global populations. Intriguingly, the genetic loci around the disease-associated SNPs showed similar LD patterns and allele frequencies around the world. The results of these analyses highlight the potential transferability of GWAS results to non-studied populations and could help inform clinical decisions in populations with a higher genetic risk of developing different complex disorders.

## Methods

### Data sets

We collected publicly available GWAS summary statistics for 20 complex disorders with no overlap with the target data. The data was cleaned to remove any duplicate and mismatched SNPs. The target dataset for the analysis consisted of 3,953 samples from 24 different countries belonging to five different ancestral groups of the globe namely, Africans (504), Europeans (2109), South Asians (489), East Asians (504) and Admixed Americans (347). The European samples were collected from previous studies (51–56) and the samples from other populations were acquired from the publicly available 1000 genomes phase 3 data (33). The detailed list or data sources are shown in supplementary table 1 and all appropriate informed consent, IRB approvals, and Data Use Agreements are in place for use of data as part of this study. The final dataset was cleaned using Plink (57) to filter out variants with more than 2% missingness, minor allele frequency <0.01 and Hardy-Weinberg Equilibrium<1e-6. After QC, we included 3,953 samples and 1,618,220 SNPs for PRS calculation.

The prevalence data for 18 traits was collected from the Global Burden of Disease (GBD) database and the prevalence information for type 2 diabetes and obesity was collected from the International Diabetes Federation (IDF) and WHO database respectively (2,58,59) (Supplementary file 4). For certain conditions like AD, CRD and CAD for which specific data isn’t available, we used the prevalence data from broad traits like dementia, IBD and ischemic heart disease. In total we gather prevalence data for 20 diseases

### Principal Component Analysis

We performed principal component analysis for both the European and Global dataset to visualize the genetic architecture of the different populations. EIGENSOFT software was used to run the analysis (60). The dataset was cleaned to remove the MHC and the chromosome 8 inversion region and LD pruned to select independent SNP to calculate the Principal Components (PCs).

### Polygenic Risk Scores Estimation

PRSice-2 was used for the estimation of PRS scores of the cleaned target dataset for each disorder with the average effect size-based scoring method (32). The tool uses Clumping and P-value Thresholding (C+T) method to select independent SNPs that are used to calculate the scores. We use a clumping threshold (r^2^) of 0.1 within a 250kb distance and 6 p-value thresholds to select the variants and calculate the scores for every individual. For the European data analyses, we clumped all the populations together, but for the global population analysis, we separated the data into five super populations as mentioned above and clumped each group separately. We then determined the standardized average PRS scores for the 24 countries that are used to visualize the distribution pattern of the various disorders and identify populations with higher genetic risk. We also used these scores to estimate the correlation between genetic risk and prevalence of a disorder.

### Correlations with prevalence and empirical p-value calculations

To determine if the average genetic risk of a disorder in a population is associated with the prevalence of disorder, we estimated Pearson’s correlations between the Average scores and the prevalence data using MATLAB R2020b. The initial level of significance was obtained using a permutation test within the data. To calculate the empirical p-value and confirm the initial significance, we performed a statistical test using a random PRS method. We first picked 100 random SNP sets to compute PRS with the number of SNPs in each set equal to the number in the significant threshold. We then computed the correlation coefficients between each random SNP set and prevalence of target disorder and determined the empirical p-value for each of the sets. We finally identified the number of SNP sets that had significant correlation higher than the PRS scores at the actual threshold and returned the empirical p-value.

### SNP Heritability

We calculated the SNP heritability of the disorders to assess whether traits with significant correlation with prevalence also have high SNP heritability. LD Score Regression (LDSC) software was used to calculate the values for each of the 20 disorders. Pre-calculated LD scores from 1000 genomes European data were used for the analysis (34).

### Linkage Disequilibrium analysis

We first start by selecting SNPs that were used for PRS calculations of various disorders in European populations, specifically at the p-value thresholds for which we observed significant correlations between the risk scores and prevalence. To determine if the regions around these SNPs are conserved across populations, we extracted all variants within a 100 KB region around the PRS SNPs and calculated r^2^ for all pairs of SNPs within the region. This was done independently for each of the five ancestral populations and repeated for all disorders separately. We then compared the r^2^ values of the various pairs of SNPs in Europeans to the values of the same pair in each of the other four populations to estimate the Pearson’s correlation for each disorder at the specific p-value threshold. To calculate an empirical p-value, we first started by making 100 SNP sets with each set having 1000 SNPs each. For each set, we then repeated the analysis as above and obtained a distribution of correlation estimates. We then used this distribution to determine if the correlations observed between Europeans and each of the other populations for different disorders are significantly higher (top 5^th^ percentile) compared to the correlation distribution obtained from the random SNP sets. The estimation of r^2^ was done using the Plink tool (57) and the statistical analyses were performed in R.

### F_ST_ Analysis

We selected SNPs that were used for PRS calculations of various disorders in European populations, at the p-value thresholds for which we observed significant correlations between the risk scores and prevalence. Then we analyzed four different groups composed of Europeans and Africans, Europeans and South Asians, Europeans and East Asians, and Europeans and admixed Americans. We calculated the F_ST_ of the selected SNPs in each group individually with each ancestry used as a sub-population and determined the mean F_ST_ of all SNPs in each pair. Analysis was repeated separately for all disorders at the specific p-value thresholds. To calculate an empirical p-value for both analyses, we created 100 sets of 1000 randomly selected SNPs and repeated the F_ST_ calculations to get a distribution. We used this distribution to verify if the mean F_ST_ of the PRS SNPs in each population pair is significantly lower (bottom 5^th^ percentile) than the distribution of the random SNP sets. The F_ST_ calculation was done using the Plink tool and the statistical analyses were performed in R.

## Data Availability

All data produced in the present study are available upon reasonable request to the authors

## Additional Files legends

**Additional file 1:** The tables show the number of SNPs used for PRS calculation of each disorder for each of the five ancestral groups. Each sheet shows the number of SNPs for each of the 6 p-value thresholds used for PRS calculation

**Additional file 2:** The tables show the Std. Average PRS Scores of each disorder in the 9 European populations. Each sheet shows the score at each of the six p-value thresholds used for PRS calculation.

**Additional file 3:** The tables show the Std. Average PRS Scores of each disorder in the 24 Global populations. Each sheet shows the score at each of the six p-value thresholds used for PRS calculation.

**Additional file 4:** The table shows the age adjusted prevalence data of the 20 disorders in 24 global populations.

