## Supplementary for "Polygenic risk scores based on European GWAS correlate to disease prevalence differences around the world"

Jain et al.

### List of Tables


1 Sources of population and sample sizes ………………………………………. 2

2 List of disorders ………………………………………………………………… 3

3 Correlations between PRS and prevalence in European populations ………. 4

4 Correlations between PRS and prevalence in Global populations…………... 5

5 SNP – Heritability Estimates ………………………………………………….. 6

6 Associations between SNP heritability and correlation estimates …………… 7

7 Correlations between $r^2$ estimates of EUR and other populations …………... 8

8 $F_{ST}$ estimates between EUR and other populations …………………………... 9


### List of Figures


1 PRS Distribution across European Populations (PRS p-value <1) ………….. 10

2 PRS Distribution across World Populations (PRS p-value <1) ……………… 11

**References** …………………………………………………………………………….. 12

**Supplementary Table 1**: Sources of the data and number of samples analyzed per population.

| Super Population | Country | Sample Size | Data Source |
|---|---|---|---|
| Europeans | Greece | 246 | Paschou et al., PNAS (1)<br>Paschou et al., Annals of Neurology (2)<br>Stamatoyannopoulos et al., EJHG (3) |
| | Italy | 192 | |
| | Hungary | 249 | |
| | Poland | 249 | TS – EUROTRAIN Study (4) |
| | Spain | 222 | |
| | Denmark | 244 | |
| | Germany | 246 | Popgen Study (5) |
| | France | 244 | Three city study(6) |
| | United Kingdom | 217 | WTCCC(code : EGAS00000000028) (7) |
| Africans | Kenya | 99 | 1000 Genomes project (8) |
| | Nigeria | 207 | |
| | Sierra Leone | 85 | |
| | Gambia | 113 | |
| South Asians | Pakistan | 96 | |
| | India | 205 | |
| | Srilanka | 102 | |
| | Bangladesh | 86 | |
| East Asians | China | 301 | |
| | Vietnam | 99 | |
| | Japan | 104 | |
| Admixed Americans | Mexico | 64 | |
| | Puerto Rico | 104 | |
| | Colombia | 94 | |
| | Peru | 85 | |

**Supplementary Table 2**: List of studied disorders and sample size of the respective GWAS studies

| Category | Disorder | N-Cases | N-Controls | References |
|---|---|---|---|---|
| **Cardiovascular** | Coronary Artery Disease (CAD) | 10,901 | 137,914 | (9) |
| **Neurological** | Alzheimer's Disease (AD) | 71,880 | 383,378 | (10) |
| | Parkinson's Disease (PD) | 33,674 | 449,056 | (11) |
| **Metabolic** | Type 2 Diabetes (T2D) | 74,124 | 824,006 | (12) |
| | Obesity (OBY) | 32,858 | 65,839 | (13) |
| | Chronic Kidney Disease (CKD) | 12,385 | 104,780 | (14) |
| | Poly Cystic Ovarian Syndrome (PCOS) | 10,074 | 103,164 | (15) |
| **Autoimmune** | Asthma (AST) | 64,538 | 329,321 | (16) |
| | Type 1 Diabetes (T1D) | 6,683 | 12,173 | (17) |
| | Psoriasis (PSO) | 10,558 | 22,806 | (18) |
| | Rheumatoid Arthritis (RA) | 14,361 | 43,923 | (19) |
| | Inflammatory Bowel Disease (IBD) | 12,882 | 21,770 | (20) |
| | Crohn's Disease (CRD) | 5,956 | 14,927 | (20) |
| | Multiple Sclerosis (MS) | 9,772 | 17,376 | (21) |
| **Psychiatric** | Attention Deficit/Hyperactivity Disorder (ADHD) | 20,183 | 35,191 | (22) |
| | Anxiety Disorder (ANX) | 7,016 | 14,745 | (23) |
| | Autism Spectrum Disorder (ASD) | 18,381 | 27,969 | (24) |
| | Bipolar Disorder (BPD) | 20,352 | 31,358 | (25) |
| | Schizophrenia (SCZ) | 69,369 | 236,642 | (26) |
| | Major Depressive Disorder (MDD) | 135,458 | 344,901 | (27) |

**Supplementary table 3:** Pearson's Correlations estimates for Average genetic risk of 20 complex disorders and their Prevalence in European populations. The column headers indicate the p-value threshold for PRS calculation and the value in each cell represents correlation coefficient and p-value based on 1000 permutations (shown in parentheses). The **(*)** indicates empirical p-value<0.05 (based on statistical test)

| Disorder | $p<5x10^{-08}$ | $p<5x10^{-05}$ | p<0.001 | p<0.01 | p<0.05 | p<1 |
|---|---|---|---|---|---|---|
| CAD | **0.72* (0.022)** | **0.62* (0.019)** | **0.74 (0.017)** | 0.51 (0.077) | 0.47 (0.137) | 0.46 (0.136) |
| PD | 0.73 (0.989) | 0.6 (0.964) | 0.41 (0.89) | 0.22 (0.333) | 0.18 (0.351) | 0.09 (0.445) |
| AD | 0.4 (0.14) | 0.3 (0.16) | 0.08 (0.43) | 0.23 (0.746) | 0.19 (0.706) | 0.05 (0.466) |
| T2D | **0.56* (0.05)** | 0.38 (0.173) | 0.21 (0.281) | 0.57 (0.065) | 0.55 (0.069) | 0.48 (0.106) |
| OBY | **0.39* (0.145)** | 0.36 (0.172) | **0.65* (0.025)** | 0.26 (0.27) | 0.08 (0.419) | 0.03 (0.508) |
| PCOS | - | 0.05 (0.502) | 0.48 (0.121) | **0.54* (0.044)** | **0.63* (0.038)** | **0.64 (0.022)** |
| CKD | 0.25 (0.249) | 0.12 (0.375) | 0.28 (0.765) | 0.31 (0.797) | 0.25 (0.737) | 0.2 (0.717) |
| AST | 0.07 (0.561) | 0.21 (0.702) | 0.27 (0.736) | 0.35 (0.847) | 0.22 (0.737) | 0.18 (0.655) |
| T1D | 0.32 (0.788) | 0.52 (0.911) | 0.46 (0.902) | 0.48 (0.914) | 0.49 (0.93) | 0.27 (0.721) |
| PSO | 0.18 (0.819) | 0.1 (0.693) | 0.28 (0.107) | 0.28 (0.1) | 0.11 (0.331) | 0.2 (0.178) |
| RA | **0.47* (0.048)** | 0.48 (0.073) | 0.53 (0.06) | 0.41 (0.111) | 0.43 (0.139) | **0.61 (0.036)** |
| IBD | 0.03 (0.488) | 0.24 (0.255) | 0.1 (0.372) | 0.1 (0.618) | 0.47 (0.877) | 0.22 (0.715) |
| CRD | 0.5 (0.921) | 0.2 (0.705) | 0.63 (0.963) | 0.11 (0.648) | 0.2 (0.689) | 0.12 (0.424) |
| MS | 0.28 (0.23) | 0.26 (0.728) | 0.25 (0.268) | 0.28 (0.243) | 0.42 (0.126) | 0.43 (0.125) |
| ADHD | 0.16 (0.604) | 0.19 (0.263) | **0.51* (0.043)** | 0.41 (0.111) | 0.05 (0.456) | 0.2 (0.294) |
| ANX | - | 0.41 (0.862) | 0.18 (0.702) | 0.54 (0.93) | 0.59 (0.954) | 0.4 (0.854) |
| ASD | 0.04 (0.611) | 0.48 (0.096) | 0.1 (0.39) | 0.06 (0.59) | 0.01 (0.457) | 0.15 (0.657) |
| BPD | 0.18 (0.723) | 0.16 (0.328) | 0.21 (0.299) | 0.13 (0.381) | 0.03 (0.535) | 0.05 (0.566) |
| SCZ | 0.38 (0.152) | **0.6* (0.041)** | **0.84* (0.002)** | **0.8* (0.006)** | **0.82 (0.009)** | **0.78 (0.009)** |
| MDD | 0.45 (0.897) | 0.2 (0.216) | 0.12 (0.613) | 0.19 (0.304) | 0.17 (0.319) | 0.23 (0.258) |

**Supplementary Table 4:** Pearson's correlations estimate for average genetic risk of 20 complex disorders and their prevalence in 24 countries. The column headers indicate the p-value threshold for PRS calculation. The value in each cell represents correlation coefficient & p-value based on 1000 permutations (shown in parentheses). The **(*)** indicates empirical p-value<0.05 (based on statistical test)

| Disorder | $p<5x10^{-08}$ | $p<5x10^{-05}$ | p<0.001 | p<0.01 | p<0.05 | p<1 |
|---|---|---|---|---|---|---|
| CAD | 0.35 (0.959) | 0.11 (0.686) | 0.31 (0.06) | 0.12 (0.68) | 0.2 (0.823) | 0.01 (0.5) |
| PD | 0.24 (0.126) | **0.39* (0.026)** | 0.61 (0.99) | 0.22 (0.85) | 0.05 (0.393) | 0.2 (0.194) |
| AD | 0.12 (0.278) | 0.22 (0.141) | 0.05 (0.292) | 0.36 (0.942) | 0.7 (1) | 0.42 (0.92) |
| T2D | 0.39 (0.957) | 0.25 (0.1) | **0.55* (0.003)** | **0.45 (0.011)** | **0.48 (0.013)** | **0.55 (0.006)** |
| OBY | **0.38* (0.039)** | 0.18 (0.805) | **0.73* (0.001)** | 0.52 (0.981) | 0.58 (1) | 0.31 (0.057) |
| PCOS | - | 0.32 (0.94) | 0.24 (0.849) | 0.27 (0.89) | 0.25 (0.862) | 0.36 (0.99) |
| CKD | 0.08 (0.7) | 0.17 (0.194) | 0.09 (0.7) | 0.08 (0.68) | 0.08 (0.631) | 0.13 (0.742) |
| AST | **0.46* (0.001)** | **0.4* (0.011)** | **0.35* (0.039)** | **0.4 (0.011)** | **0.32 (0.048)** | 0.55 (1) |
| T1D | **0.42* (0.009)** | **0.39* (0.028)** | **0.41* (0.022)** | 0.31 (0.076) | **0.42 (0.022)** | 0.18 (0.226) |
| PSO | 0.18 (0.819) | 0.1 (0.693) | 0.28 (0.107) | 0.28 (0.1) | 0.11 (0.331) | 0.2 (0.178) |
| RA | 0.05 (0.402) | **0.58* (0.004)** | 0.14 (0.293) | 0.08 (0.35) | 0.32 (0.949) | 0.39 (0.969) |
| IBD | **0.41* (0.017)** | **0.33* (0.007)** | **0.34* (0.05)** | 0.39 (0.971) | 0.25 (0.879) | 0.03 (0.429) |
| CRD | 0.4 (0.985) | 0.44 (0.98) | 0.32 (0.06) | **0.6* (0.001)** | **0.55 (0.001)** | **0.38 (0.035)** |
| MS | **0.42* (0.014)** | 0.19 (0.183) | 0.09 (0.645) | **0.49 (0.001)** | **0.49 (0.001)** | **0.71 (0.001)** |
| ADHD | 0.05 (0.586) | **0.46* (0.008)** | 0.12 (0.333) | 0.09 (0.386) | **0.38 (0.014)** | 0.26 (0.096) |
| ANX | - | 0.14 (0.764) | 0.01 (0.519) | **0.5* (0.002)** | 0.22 (0.015) | 0.04 (0.557) |
| ASD | 0.15 (0.244) | 0.02 (0.535) | 0.18 (0.22) | 0.03 (0.464) | 0.04 (0.423) | 0.08 (0.361) |
| BPD | 0.28 (0.1) | **0.42* (0.013)** | **0.49* (0.004)** | 0.15 (0.763) | 0.28 (0.863) | 0.07 (0.376) |
| SCZ | **0.71* (0.001)** | **0.45* (0.011)** | 0.141 (0.723) | 0.32 (0.967) | 0.18 (0.784) | 0.19 (0.798) |
| MDD | 0.27 (0.89) | 0.2 (0.81) | 0.32 (0.918) | 0.39 (0.97) | 0.07 (0.62) | 0.01 (0.47) |

**Supplementary table 5:** SNP – Heritability estimates of 20 complex disorders calculated using LDSC using GWAS summary statistics.

| Disorder | SNP - Heritability |
|---|---|
| Coronary Artery Disease (CAD) | 0.099 |
| Alzheimer’s Disease (AD) | 0.0145 |
| Parkinson’s Disease (PD) | 0.0113 |
| Type 2 Diabetes (T2D) | 0.0286 |
| Obesity (OBY) | 0.1547 |
| Chronic Kidney Disease (CKD) | 0.0246 |
| Poly Cystic Ovarian Syndrome (PCOS) | 0.044 |
| Asthma (AST) | 0.075 |
| Type 1 Diabetes (T1D) | NA |
| Psoriasis (PSO) | 0.5225 |
| Rheumatoid Arthritis (RA) | 0.143 |
| Inflammatory Bowel Disease (IBD) | 0.537 |
| Crohn’s Disease (CRD) | 0.86 |
| Multiple Sclerosis (MS) | 0.0492 |
| Attention Deficit/Hyperactivity Disorder (ADHD) | 0.2354 |
| Anxiety Disorder (ANX) | 0.0768 |
| Autism Spectrum Disorder (ASD) | 0.16 |
| Bipolar Disorder (BPD) | 0.3 |
| Schizophrenia (SCZ) | 0.157 |
| Major Depressive Disorder (MDD) | 0.0214 |

**Supplementary table 6:** Associations between SNP Heritability and the correlation estimates of PRS-Prevalence of 20 disorders at different p-value thresholds

| P-value threshold | Correlation estimate | p-value |
|---|---|---|
| 5e-08 | 0.12 | 0.65 |
| 5e-05 | 0.21 | 0.39 |
| 0.001 | 0.10 | 0.70 |
| 0.01 | 0.24 | 0.33 |
| 0.05 | 0.13 | 0.60 |
| 1 | -0.13 | 0.59 |

**Supplementary table 7**: Correlations between $r^2$ estimates of SNP pairs in regions used for PRS estimation in Europeans to other populations for disorders with significant correlations. The significant results (Empirical p-value <0.05) are indicated as bold.

| **Disorder – pvalue** | **EUR – AFR** | **EUR – SAS** | **EUR – EAS** | **EUR – AMR** |
|---|---|---|---|---|
| Mean (Random Set) | 0.7894413254 | 0.9623010762 | 0.8971119236 | 0.9634032023 |
| PD-5E-05 | **0.8186459379** | 0.9574071602 | 0.8853407419 | **0.9683809978** |
| T2D-0.001 | 0.791407982 | 0.9628390958 | 0.8976396671 | **0.9656916419** |
| OBY-5E-08 | **0.8114106098** | **0.9738592567** | **0.9332260314** | **0.9764401093** |
| OBY-0.001 | **0.7959332968** | **0.965722474** | 0.8978775434 | **0.9689971174** |
| AST-5E-08 | 0.7684976549 | 0.9595141279 | 0.8812201287 | **0.9672762241** |
| AST-5E-05 | 0.7777785261 | 0.9586197328 | 0.8843342743 | **0.9631166733** |
| AST-0.001 | 0.7920626098 | 0.9624762676 | 0.89493203 | **0.9646682301** |
| T1D-5E-08 | **0.8150671692** | **0.9647759291** | 0.8349460058 | 0.9417422395 |
| T1D-5E-05 | **0.8032053635** | **0.9694990408** | 0.8798865479 | 0.9520407467 |
| T1D-0.001 | 0.7916303711 | **0.9644417155** | 0.882218677 | 0.9552061935 |
| RA-5E-05 | 0.7857483628 | 0.9633299006 | **0.9056577697** | **0.9714361793** |
| IBD-5E-08 | **0.7982407945** | 0.9631529015 | 0.8988146243 | **0.9694391631** |
| IBD-5E-05 | **0.797444013** | **0.9644363861** | 0.8977089739 | **0.9659353958** |
| IBD-0.001 | **0.8008613501** | **0.9651375325** | 0.8986818172 | **0.9672679062** |
| CRD-0.01 | **0.8005422308** | 0.9631338031 | 0.8962575684 | **0.9660411078** |
| MS-5E-08 | **0.8039129249** | **0.9676498263** | **0.9214971578** | **0.9706299851** |
| ADHD-5E-05 | **0.8032160313** | **0.9647792732** | 0.8854770614 | **0.9658057354** |
| ANX-0.01 | **0.8017354755** | 0.9618246591 | 0.8960207127 | **0.965050267** |
| BPD-5E-05 | 0.7922255933 | 0.9630197803 | 0.8977957091 | **0.9648965738** |
| BPD-0.001 | 0.7926035047 | 0.9630635376 | 0.8951110061 | 0.9631121371 |
| SCZ-5E-08 | 0.7657702533 | **0.970065904** | 0.8962770909 | **0.9657351075** |
| SCZ-5E-05 | 0.7855139566 | **0.9648879414** | 0.8933117915 | **0.9646781049** |

**Supplementary table 8**: Mean $F_{ST}$ estimates of PRS SNPs between EUR and other populations for disorders with significant correlations. The significant results (empirical p-value < 0.05) are indicated as bold.

| **Disorder – pvalue** | **EUR – AFR** | **EUR – SAS** | **EUR – EAS** | **EUR – AMR** |
|---|---|---|---|---|
| Mean (Random set) | 0.108265 | 0.0334789 | 0.08526447 | 0.0249905 |
| PD-5E-05 | 0.122477 | 0.0498318 | 0.113105 | 0.0333909 |
| T2D-0.001 | 0.105748 | 0.0320757 | 0.0862362 | 0.0241245 |
| OBY-5E-08 | **0.0998567** | **0.0211867** | 0.104279 | 0.0275066 |
| OBY-0.001 | 0.109003 | 0.0299368 | 0.087189 | **0.0229615** |
| AST-5E-08 | **0.0932677** | 0.0323666 | 0.0796585 | **0.0231094** |
| AST-5E-05 | **0.0995356** | 0.0308866 | **0.0789205** | 0.0248309 |
| AST-0.001 | **0.0967675** | 0.0323278 | 0.0813934 | 0.0242087 |
| T1D-5E-08 | 0.141237 | **0.0287936** | 0.161305 | 0.0318749 |
| T1D-5E-05 | 0.0987654 | 0.031684 | 0.117239 | 0.0238838 |
| T1D-0.001 | 0.111469 | **0.0293318** | 0.0924458 | **0.021092** |
| RA-5E-05 | **0.0996238** | 0.0383163 | 0.084651 | **0.021027** |
| IBD-5E-08 | **0.101194** | 0.0369472 | 0.0882059 | **0.0204246** |
| IBD-5E-05 | 0.103456 | 0.0341029 | 0.0817805 | **0.0216712** |
| IBD-0.001 | 0.103485 | 0.0333184 | 0.0856894 | 0.0230368 |
| CRD-0.01 | **0.103137** | 0.0311598 | 0.0821845 | 0.0239542 |
| MS-5E-08 | **0.0905286** | 0.0506884 | 0.115611 | 0.0316858 |
| ADHD-5e-05 | 0.10517 | 0.0466617 | 0.0836218 | 0.0249051 |
| ANX-0.01 | **0.0966324** | 0.0330105 | 0.0848371 | 0.0253049 |
| BPD-5E-05 | **0.101407** | 0.0315619 | 0.0887759 | 0.0260013 |
| BPD-0.001 | 0.104078 | 0.0326655 | 0.0850312 | 0.0249228 |
| SCZ-5E-08 | 0.137028 | 0.0325683 | 0.0992565 | 0.0254695 |
| SCZ-5E-05 | 0.113266 | 0.0315388 | 0.0916087 | 0.0234403 |

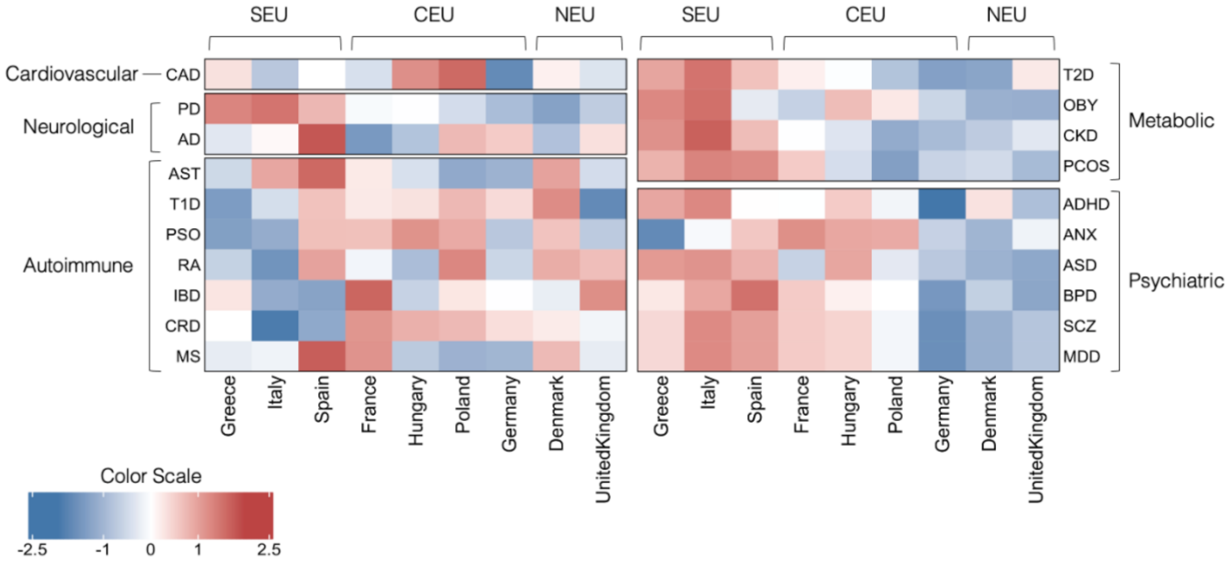


**Supplementary Figure 1: Heatmap of Std. Average PRS ($r^2$ = 0.1; p-value<1) of 20 Disorders across European Populations**. Populations are arranged based on their geographical proximities. Shades of cells indicate the standardized avg. genetic risk of each disorder for each population. A higher risk is shown by red and lower risk is indicated by blue [SEU – South Europeans, CEU – Central Europeans, NEU – North Europeans.]

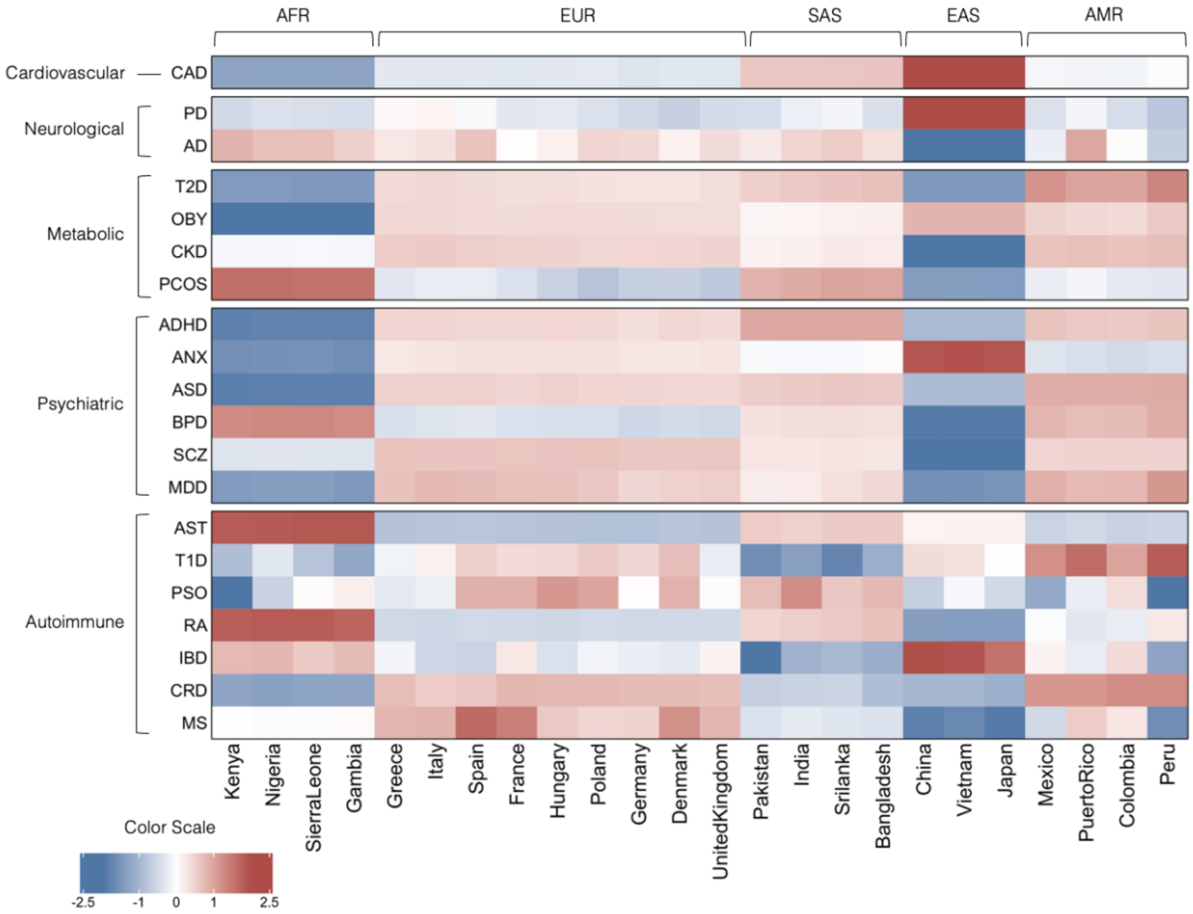


**Supplementary Figure 2: Heatmap of Std. Average PRS (p-value<1) of 20 Disorders across Worldwide Populations**. Populations are arranged based on their geographical locations and ancestry. Shades of cells indicate the standardized avg. genetic risk of each disorder for each population. A higher risk is shown by red and lower risk is indicated by blue. [AFR – Africans, EUR – Europeans, SAS – South Asians, EAS – East Asians, AMR – Admixed Americans]
